# Using polygenic and transcriptional risk scores to investigate psychiatric and cognitive symptoms in Parkinson’s disease

**DOI:** 10.1101/2025.03.20.25324166

**Authors:** Lachlan Gilchrist, Oliver Pain, Heather Marriott, Abigail Pfaff, Cameron James Watson, Alfredo Iacoangeli, Sulev Koks, Cathryn M. Lewis, Petroula Proitsi

**Affiliations:** Social, Genetic and Developmental Psychiatry Centre, Institute of Psychiatry, Psychology, and Neuroscience, King’s College London, London, United Kingdom; Perron Institute for Neurological and Translational Science, Perth, Australia; Maurice Wohl Clinical Neuroscience Institute, Department of Basic and Clinical Neuroscience, Institute of Psychiatry, Psychology and Neuroscience, King’s College London, London, United Kingdom; Department of Biostatistics and Health Informatics, Institute of Psychiatry, Psychology and Neuroscience, King’s College London, London, United Kingdom; Personalised Medicine Centre, Health Futures Institute, Murdoch University, Perth, Australia; South London and Maudsley NHS Foundation Trust, London, United Kingdom; NIHR Maudsley Biomedical Research Centre at South London and Maudsley NHS Foundation Trust and King’s College London, King’s College London, London, United Kingdom; Department of Medical and Molecular Genetics, King’s College London, London, United Kingdom; Centre for Preventive Neurology, Wolfson Institute of Population Health, Queen Mary University of London

**Author notes:** Joint senior corresponding authors: Dr. Petroula Proitsi; Prof. Cathryn M. Lewis; Prof. Sulev Kõks.

## Abstract

Psychiatric and cognitive symptoms are commonly observed in individuals with Parkinson’s disease (PD) with a significant impact on quality of life. Given the genetic contribution to PD, cross-trait polygenic risk score (PRS) analysis may elucidate underlying biological mechanisms. Transcriptional risk scores (TRS) – the weighted sum of an individual’s observed gene expression – offer a complementary approach, having shown good prediction in other diseases. Here, we combined transcriptome-wide analysis of Alzheimer’s disease (AD), depression, PD, and schizophrenia with RNA-seq data from the Parkinson’s Progressive Markers Initiative (PPMI) (*N* = 592; *N_CASES_* = 431) and 10 years of follow-up data to conduct the first within- and cross-trait TRS study of PD, its severity, psychiatric and cognitive symptoms, and their progression. TRS associations were compared to PRS. Polygenic risk for PD was significantly associated (*p_FDR_* < 0.05) with PD case/control status. Higher transcriptional risk for PD was not associated with PD case/control status but was associated with greater PD severity at baseline, suggesting they still capture the broader disease burden. Higher transcriptional risk for AD was associated with having PD, greater baseline severity, higher baseline depression, anxiety, hallucinations and a faster increase in both hallucinations and apathy, implicating shared mechanisms across neurodegenerative diseases. Higher polygenic and transcriptional risk for depression was associated with higher baseline depression and anxiety in PD and a faster increase in anxiety over time. Higher transcriptional risk for AD was associated with faster decline across all cognitive domains, while higher transcriptional risk for schizophrenia showed protective effects, suggesting TRS can identify novel biological contributions to cognitive decline, with the potential for targeted interventions. Our findings show that transcriptional and polygenic risk scores capture different aspects of biological contributions to PD symptoms and progression.

## 1. Introduction

Although Parkinson’s disease (PD) is clinically characterised by its motor symptoms^1^, cognitive and psychiatric symptoms are common. Individuals with PD are at approximately six times greater risk of dementia compared to non-PD individuals of a similar age^2^ and 20-50% will experience cognitive impairment during the disease course^3^. Further, anxiety, apathy, depression and psychosis are observed in between 30-40% of patients^4^, with comorbid PD depression and psychosis associated with faster motor progression and cognitive decline^5^. These non-motor symptoms have significant impact on the quality of life of individuals with PD and increase carer burden^2,6^. To improve long-term outcomes for individuals with PD, developing a deeper understanding of the risk mechanisms underlying cognitive and psychiatric symptoms is vital.

Although several genes – such as *GBA1*, *LRRK2*, *PRKN* and *SNCA* – are considered high risk or monogenic in their contribution to PD^7^, a polygenic contribution is well evidenced. In genome-wide association analysis (GWAS), 90 independent risk loci have been identified in individuals of European ancestry, with approximately 22% of the phenotypic variance explained by common variants^8^. GWAS may be leveraged for disease prediction through polygenic risk scores (PRS) – the sum of an individual’s risk alleles weighted by that allele’s GWAS effect size^9^. PRS can also be used to infer the effects of genetic risk for one phenotype on another, allowing us to better understand shared biology^10^. Such cross-trait analyses as applied within PD have shown that higher genetic risk for PD is associated with an earlier age of onset^11^ and faster cognitive and motor decline^12^. Additionally, higher genetic risk for Alzheimer’s disease (AD) is associated with hallucinations in PD, driven primarily by the effects of the Apolipoprotein E (*APOE*) region^13^ – a locus whose associations with neurodegeneration are well-documented across brain diseases^14^.

However, the variance explained by PD-PRS remains modest at just over 5% for PD case/control status on the liability scale^8^. Transcriptional risk scores (TRS) – the weighted sum of an individual’s gene expression – offer a complementary approach, having shown predictive utility in studies of attention-deficit/hyperactivity disorder (ADHD)^15^, amyotrophic lateral sclerosis (ALS)^16^, AD^17,18^ and Crohn’s disease^19^. However, this approach has yet to be applied to PD despite ∼2000 genes being differentially expressed in PD patients compared to controls^20^. Further, this approach has yet to applied cross-trait to infer the associations between the gene expression profile of one disease on another.

As such, in this study, we calculate PRS/TRS for AD, depression, PD and schizophrenia in the Parkinson’s Progressive Markers Initiative (PPMI)^21,22^. We assess the association between PRS/TRS and (1) PD-specific phenotypes (PD case/control status, age of onset and overall disease severity); (2) psychiatric symptoms (depression, anxiety, apathy and hallucinations); and (3) cognitive outcomes (processing speed, visuospatial ability, working memory, verbal learning, and overall cognitive impairment). Further, we extract individual-level slopes for overall disease severity and psychiatric and cognitive trajectories from up to 10 years of follow-up to assess the effect of PRS/TRS on their rate of change.

## 2. Methods

### 2.1 Study sample

The Parkinson’s Progressive Markers Initiative (PPMI) is a global, multi-centre observational study launched in 2010 with the goal of identifying novel biomarkers of PD progression^21^. PD cases enrolled in the study were ≥30 years old and within two years of clinical diagnosis, scoring ≤3 on the Hoehn and Yahr scale and presenting at least two of bradykinesia, resting tremor, or rigidity. All cases in the original cohort were required to be unmedicated at recruitment. Healthy controls were recruited with the same age criteria and required to be without a current neurological disorder and no PD diagnosis among immediate family.

The PPMI data cut from the consensus committee analytic dataset (release: 12/06/2023) was obtained from the Laboratory of Neuro Imaging Image Data Archive (LONI IDA) (*N* = 2,347). Participants classified as prodromal for PD were not included in our analyses (*N* = 958). Individuals were further filtered to exclude those self-reporting non-European ancestry (*N* = 162) and those without both RNA-seq and whole genome sequencing data (*N* = 577). Within the GenoPred pipeline v.2.2^23^ – a reference standardised framework for polygenic scoring – related individuals were excluded using PLINK v2^24^ and a KING relatedness threshold > 0.044 (third-degree relatives) (*N* = 4). Further, individuals of non-European genetic ancestry were identified and excluded using principal component analysis and the combined 1000 Genomes Phase 3 (1KG)^25^ and Human Genome Diversity Project (HGDP)^26^ reference population sample (*N* = 3313), assigned to ancestry clusters with a predicted probability threshold of ≥90%. The final sample contained 431 cases and 161 healthy controls for case/control analyses. For all other outcomes, analyses were restricted to PD cases only (*N_MAX_* = 431). We compared the age and sex of cases and controls using a t-test and chi-square test.

### 2.2 Study outcomes

We assessed the predictive utility of the PRS/TRS on PD case/control status, age of onset (*N* = 424) and overall PD severity, measured by each individual’s total score on the Movement Disorder Society Unified Parkinson’s Disease Rating Scale (MDS-UPRS) (OFF medication)^27^ (*N* = 400).

For psychiatric outcomes, depression was assessed on the Geriatric Depression Scale (GDS)^28^ (*N* = 430) and anxiety on the State-Trait Anxiety Inventory (STAI)^29^ (*N* = 430). For apathy and hallucinations, we used two items from part one of the MDS-UPRS measuring apathy and hallucination severity. Both items measure the severity of symptoms on a five-point scale (0-4) where a score of zero represents no presence of that symptom over the past week and scores one to four represent severity as slight, mild, moderate and severe. Due to difficulty interpreting ordinal variables and the low number of individuals with high severity values we transformed these into binary phenotypes with scores of one to four defined as cases. Case numbers for hallucinations at baseline remained low (*N_CASES_* = 16; *N_CONTROLS_* = 413), with slightly more cases available for apathy (*N_CASES_* = 73; *N_CONTROLS_* = 356).

We additionally assessed the association between the PRS/TRS and four individual cognitive domains: information processing speed (Symbol Digit Modality test^30^) (*N* = 429), visuospatial ability (Benton Judgment of Line Orientation^31^) (*N* = 428), working memory (Letter-Number Sequencing test^32^) (*N* = 428) and verbal learning (Hopkins Verbal Learning Test total score^33^) (*N* = 429). Associations with global cognitive impairment were assessed using scores from the Montreal Cognitive Assessment^34^ (MoCA) (*N* = 431). MoCA scores used in this study were pre-adjusted for educational attainment by PPMI investigators prior to access.

#### 2.2.4 Longitudinal slopes

For overall PD severity and all cognitive and psychiatric measures, we used linear mixed-effects models with random intercept and slopes to extract individual-level slopes for use as downstream outcomes representing the rate of change in each phenotype, as per previous analyses of cognitive trajectories^35,36^. Slopes were calculated using lme4 v.1.1.35.5, controlling for the fixed effects of sex and age at baseline, with the individual as the random effect over time. Individuals with only baseline values were excluded from the dataset (*N* = 19). Individuals were further excluded on a per-outcome basis if they had less than two total time points with non-missing data (including baseline), using up to 10 years of follow-up data. This resulted in a maximum of 5 further exclusions across the outcomes, except for overall PD severity (total OFF medication MDS-UPRS score), where 49 additional individuals were excluded (Supplementary Table 1). The mean number of non-missing follow-up timepoints per individual was greater than seven for all variables except overall PD severity, which had a mean value of six (Supplementary Table 2).

### 2.3 Genetic data

Genotype data was obtained from the PPMI whole genome sequenced (WGS) variant call format (VCF) files. PPMI samples were sequenced using Illumina HiSeq X Ten Sequencer and paired-end 300–400bp reads, processed using the Broad’s functional equivalence pipeline^37^ and aligned to the GRCh38DH reference genome, with single nucleotide polymorphisms (SNPs) called using GATK^38^. PPMI WGS data has a mean coverage of 34.2x and a mean read depth of 35.7x^39^. Further details on PPMI WGS data can be found on the LONI IDA web portal (https://ida.loni.usc.edu/pages/access/geneticData.jsp) and is described in the paper from Accelerating Medicines Partnership – Parkinson’s Disease (AMP-PD)^39^. Using *BCFTools*^40^ we extracted the 1,204,449 HapMap3 variants used by the GenoPred pipeline^23^ (see below) from the VCFs and converted them to PLINK binary format using PLINK v1.9^41^.

### 2.4 RNA-seq

PPMI gene-level RNA abundance was obtained from LONI IDA, originally derived from whole blood samples sequenced on the Illumina NovaSeq 6000 platform in lengths of 125 nucleotides at ∼100 million paired reads per sample. The FASTQ files had been aligned using STAR v.2.6.1d to GRCh38 and transcript abundance quantified with Salmon v.0.11.3 using GENCODE v.29. Further detail on the quality control process relating to this RNA-seq data can be viewed in the original paper^20^. Prior to TRS calculation, we excluded genes with low expression values using the default setting of the *filterByExpr* function of EdgeR v.3.36.0 leaving 23,298 genes. Trimmed-Mean of M-values (TMM) normalisation was applied to account for library size differences. Finally, gene expression values were log2 transformed and standardised (mean = 0; SD = 1).

### 2.5 GWAS summary statistics

For AD we used a subset from Wightman et al.^42^ (*N* = 398,058, *N_CASES_* = 39,918), excluding cases/controls ascertained based on family history (proxy)^43^ as their inclusion has been demonstrated to bias downstream genetic analyses^44,45^, as well as samples from 23andMe. For depression we obtained summary statistics from the Psychiatric Genomics Consortium (PGC)^46^ (*N* = 500,199, *N_CASES_* = 170,756), Million Veteran Program^47,48^ (*N* = 250,215; *N_CASES_* = 83,810), and FinnGen freeze 9 (F5_DEPRESSIO)^49^ (*N* = 372,472; *N_CASES_* = 43,280) and performed inverse-variance-weighted genome-wide meta-analysis using METAL^50^ for a total sample size of 1,122,886 (*N_CASES_* = 298,038). Schizophrenia summary statistics were obtained from the latest GWAS from the PGC^51^ (*N* = 130,644, *N_CASES_* = 53,386) and also meta-analysed with FinnGen freeze 9^49^ (KRA_PSY_SCHIZODEL) (*N* = 290,587, *N_CASES_* = 13,061) for a total sample size of 421,231 (*N_CASES_* = 66,447). To avoid sample overlap between base and target dataset, which can lead to inflation and false-positives^52^, for PD we used the combined UK Biobank + FinnGen + Million Veteran Program meta-analysis made publicly available by FinnGen investigators (https://mvp-ukbb.finngen.fi) (*N* = 1,533,695; *N_CASES_* = 17,624). Although this is a multi-ancestry GWAS, PRS generated from multi-ancestry base datasets show equivalent or better performance in European target samples^53,54^. All summary statistics were quality controlled prior to any analysis using MungeSumstats^55^ with dbsnp 144 and the BSgenome.Hsapiens.1000genomes.hs37d5 reference genome. Duplicate, non-biallelic and rare (MAF < 0.01) variants were excluded, and effects and allele frequencies aligned to the reference genome.

### 2.6 Polygenic risk scores (PRS)

SNP weighting was conducted within the GenoPred pipeline^23^ according to the MegaPRS^56^ method from the LDAK software. MegaPRS assumes a BLD-LDAK heritability model, incorporating information on a range of functional annotations – such as CpG content and recombination rate – alongside linkage disequilibrium (LD) and allele frequency to maximise prediction. It does not require a tuning sample and performs well in comparison to other PRS methods^57^.

### 2.7 TWAS gene identification

Gene weights for the TRS were derived using transcriptome-wide association analysis (TWAS) in FUSION^58^. FUSION allows users to estimate the predicted effect of gene expression on a phenotype of interest using the cis-SNP z-scores from GWAS summary statistics and pre-computed SNP weights based on SNP-gene expression correlations in an eQTL reference panel. The 1000 Genomes phase 3 LD reference panel required to run FUSION was obtained from the FUSION website (https://data.broadinstitute.org/alkesgroup/FUSION/LDREF.tar.bz2) (*N* = 489).

Although only whole blood RNA-seq data is available for individuals in PPMI, several studies have shown that using TWAS weights from brain eQTL panels in blood-based TRS calculation improves prediction^15,16^. As such, we identified gene expression effects using weights from three blood (*N_SAMPLE_* range = 670-1,264) and fourteen brain (*N_SAMPLE_* range = 100-452) eQTL reference panels (Supplementary Table 3), available from the FUSION website (http://gusevlab.org/projects/fusion/#single-tissue-gene-expression). For TWAS analysis, a Bonferroni-adjusted significance threshold of 6.5 x 10^-7^ was calculated using the mean number of gene-tissue TWAS tests performed across all four traits (0.05/(307,208/4)), corresponding to a *z*-score of +/- 4.976 with a two-tailed test. We applied statistical colocalisation using the COLOC^59^ extension of FUSION with default priors (p1 = 1 x 10^-4^, p2 = 1 x 10^-4^, p12 1 x 10^-5^) to identify TWAS associations with evidence of a causal variant shared between gene expression and phenotype. A posterior probability of ≥ 0.8 for H_4_ (a shared causal variant) was taken as evidence of colocalisation. TWAS results from each panel were concatenated for each phenotype prior to TRS calculation.

### 2.8 Transcriptional risk score (TRS) calculation

A TRS represents the sum of an individual’s normalised gene expression values weighted by their TWAS z-score. As per previous work^16^, we calculated a predicted gene expression matrix using genotypes from the European-ancestry sample of the 1000 Genomes Project phase 3 (*N* = 503) and applied a clumping and thresholding approach to exclude TWAS features correlated at an *R^2^* of 0.1 within +/-250kb for lead genes using the IFRisk script (https://github.com/opain/Inferred-functional-risk-scoring). Clumping was performed at *p*-value thresholds (pT) of 0.1, 0.05, 0.001, 1 x 10^-4^, 1 x 10^-6^ and the above TWAS-derived Bonferroni threshold (6.5 x 10^-7^). This approach is analogous to the clumping and thresholding approach implemented in the PRS software PRSice-2^60^. Where the same TWAS gene was retained after clumping across multiple tissues, we used their mean *z*-score to obtain a single weight for that gene as in previous work^16^. Given the complex LD structure of the major histocompatibility complex (MHC) (GRCh37: chr6:28,477,797-33,448,354), only the top gene from this region was retained during clumping. TRS were additionally calculated across *p*-value thresholds including only colocalising genes (*PP.H_4_* ≥ 0.8), and these scores were included when identifing the best performing TRS score for each outcome. Whether the best-performing TRS contained only colocalising genes or not is indicated alongside the reported *p*-value clumping threshold (pT_COLOC_ or pT, respectively).

### 2.8 Statistical analyses

Analyses were conducted using R v.4.3.1. All baseline PRS and TRS regression models adjusted for age and sex, except for PD age-of-onset (no age adjustment). For the slope outcomes, age and sex was already included in the mixed effect model. All PRS models controlled for the first 10 principal components (PCs) – calculated in GenoPred – to mitigate the effects of population stratification^61^. To account for hidden confounding in our TRS regression models – such as batch effects – we performed surrogate variable analysis (SVA) with the ‘leek’ method^62,63^ on the gene expression matrix. During SVA we protected disease status, age and sex as previously described^19^. SVA identified two surrogate variables (SVs), which were included in each TRS regression model, akin to PC inclusion for PRS.

We used linear regression to assess the predictive ability of the PRS/TRS on PD case/control status, converting *R^2^* values to the liability scale using Lee’s formula^64^ with a population prevalence of 1%. Logistic regression was used for the case/control analysis of apathy and hallucinations. We report Nagelkerke’s pseudo-*R^2^*for apathy and hallucinations. All other outcomes were assessed using linear regression, with observed *R^2^* reported. Depression and anxiety scores were log transformed prior to analysis. To improve interpretability of results, PRS/TRS and continuous outcomes were standardised (mean = 0, SD = 1) so that effect sizes represented standard deviation change in the outcome per standard deviation change in the PRS/TRS.

### 2.9 Multiple testing correction

As per previous work^15^, we use a permutation test to control for TRS type 1 error, calculating an empirical *p*-value for each TRS score for each outcome if nominally associated at *p* < 0.05. To do this, we permute the outcome 10,000 times and repeat the regression analysis. The empirical *p*-value is derived from the sum of permuted *p*-values less than the observed *p*-value plus one, divided by the total number of permutations plus one, as per the formula from Choi et al. in PRSice-2^60^. We then perform false discovery rate correction (FDR)^65^ across all PRS and the empirical *p*-values from the best performing TRS (*N_TESTS_* = 176), defining statistical significance as *p_FDR_* < 0.05.

### 2.10 Sensitivity analyses

For FDR significant associations, we performed a series of covariate-adjusted sensitivity analyses to assess potential confounding, controlling separately and in a full model for body mass index, years of education (EduYears) (except MoCA), the number of APOE ε4 risk variants (0, 1 or 2), and levodopa equivalent dose (LEDD).

As the *APOE* region (chr19:45,020,859–45,844,508 (GRCh37)) has well-documented effects on AD risk and cognitive decline,^66^ we perform additional sensitivity analyses for any statistically significant AD-PRS/TRS associations to test whether they were driven primarily by the effect of this region, excluding genes with overlapping boundaries in this region from the TRS or excluding variants in this region from the PRS.

## 3 Results

### 3.1 Descriptive statistics

No significant differences were observed between cases (*N* = 431) and controls (*N* = 161) for age (mean age [SD] = 62.216 [9.348]) vs. 61.684 [10.85], respectively) (*p* = 0.583), sex (% female = 39.907% vs. 34.782%, respectively) (*p* = 0.296), BMI (mean BMI [SD] = 26.917 [4.604] vs. 26.94 [4.489], respectively) (*p* = 0.955) and years of education (mean years of education [SD] = 15.582 [3.211] vs. 16.112 [2.945] (*p* = 0.059). The number of APOE ε4 risk variants (Supplementary Table 4) was not associated with PD case/control status in an unadjusted logistic regression (OR [95%CI] = 0.996 [0.922 - 1.069], *p* = 0.845).

PD cases had a mean age of onset of 59.59 (SD = 9.67). LEDD was non-zero for 23.90% of cases (*N* = 103) (Mean [SD] = 126.75 [287.35]). Means and standard deviations for baseline measures in PD cases can be seen in Supplementary Table 5. For slope values extracted from the mixed effects models for cases, slopes declined on average for all cognitive outcomes (mean slope range: -0.854 - -0.076) while for psychiatric outcomes and overall PD severity slopes increased (mean slope range: 0.038 - 3.799) (Supplementary Table 6). As such, negative associations between PRS/TRS and cognitive slopes indicate faster decline and positive associations slower decline, whereas negative associations with psychiatric and severity slopes indicate slower increase and positive associations faster increase.

### 3.2 TWAS gene identification

TWAS with FUSION identified a total of 2,131 gene-tissue associations after Bonferroni correction (p ≤ 6.5 x 10^-7^) across the four neuropsychiatric disorders (*N_AD_* = 163; *N_DEP_* = 508; *N_PD_* = 173; *N_SCZ_* = 1287), involving 1895 unique gene-tissue combinations and 664 unique genes. Of these, 896 associations showed evidence of colocalisation (PP.H_4_ ≥ 0.8) (*N_AD_* = 81; *N_DEP_* = 213; *N_PD_* = 100; *N_SC_*_Z_ = 502), involving 786 unique gene-tissue combinations and 319 unique genes. As the purpose of this study is not gene identification, a full table of Bonferroni significant TWAS associations can be seen in Supplementary Table 7.

### 3.3 Statistical analyses

The number of genes in final TRS scores at each clumping threshold can be seen in Supplementary Table 8. Associations between all outcomes and all PRS and the best performing TRS can be seen in Supplementary Table 9, with associations with all TRS at all clumping thresholds in Supplementary Table 10.

#### 3.3.1 PD case/control status, age of onset and disease severity

Polygenic and transcriptional risk score associations with PD case/control status, age of onset and both baseline and longitudinal PD severity can be seen in Figure 1a.

**Figure 1:**
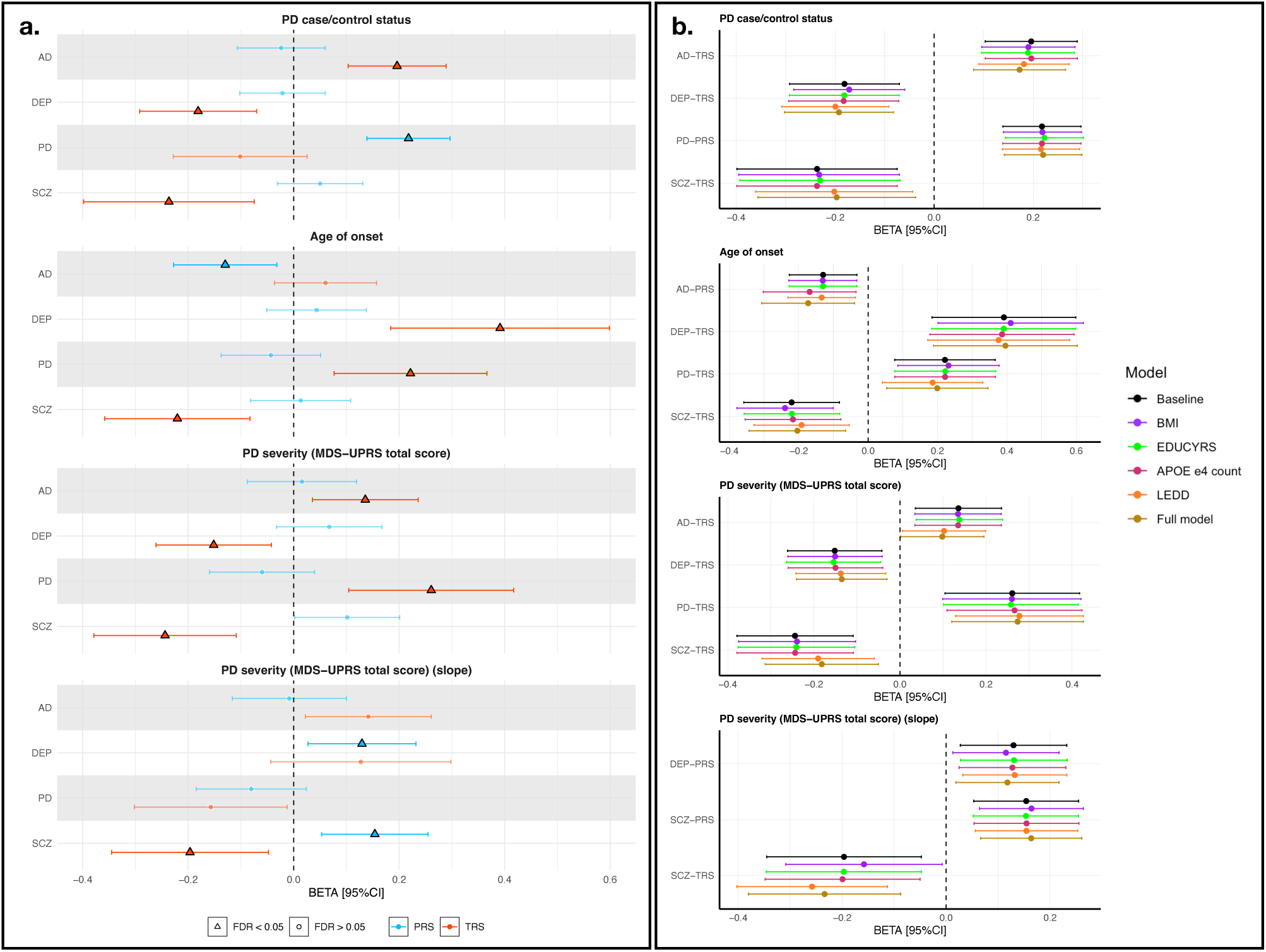
Forest plot for associations between Alzheimer’s disease (AD), depression (DEP), Parkinson’s disease (PD) and schizophrenia (SCZ) polygenic/transcriptional risk and PD case/control status, PD age of onset, overall PD severity as measured by the MDS-UPRS total score, and the longitudinal slope of PD severity. (a) Primary analysis. Point estimates for associations with p_FDR_ < 0.05 are highlighted as triangles with a black outline. Whether the estimate is from polygenic (PRS) or transcriptional risk scores (TRS) is indicated in blue or orange, respectively. (b) Covariate-adjusted sensitivity analyses for estimates with p_FDR_ < 0.05. Baseline models (e.g unadjusted models) are indicated in black, models adjusting for body mass index (BMI) in purple, years of education (EDUCYRS) in green, number of APOE e4 variants in pink, levodopa equivalent dose (LEDD) in yellow and fully adjusted models, adjusted for all covariates, in gold.

Polygenic risk for PD was significantly associated with PD case/control status after FDR correction (*R^2^_LIABILITY_* = 0.032, OR [95%CI] = 1.243 [1.149-1.345], *p_FDR_* = 1.568 x 10^-5^). By contrast, no significant association was observed for PD transcriptional risk even at a nominal significance level (*p* > 0.05). However, higher transcriptional risk for AD was significantly associated with PD case/control status (pT_COLOC_ = 1 x 10^-4^, *R^2^_LIABILITY_* = 0.0196, OR [95%CI] = 1.217 [1.109-1.335], *p_FDR_* = 5.866 x 10^-3^). No genes included in the AD-TRS overlapped with the *APOE* gene boundaries, indicating effects were not driven primarily by the expression of genes in this region. Higher transcriptional risk for depression and schizophrenia were both associated with lower odds of PD (pT_DEP_ = Bonferroni; pT_SCZ_ = 0.001, *R^2^_LIABILITY_* range = 0.009-0.012, OR range = 0.789-0.834 *p_FDR_* = 0.02-0.031).

Higher polygenic risk for AD was associated with an earlier age of PD onset (*R^2^* = 0.015, *β* [95%CI] = -0.13 [-0.227---0.032], *p_FDR_* = 0.039). Exclusion of the *APOE* region from the AD-PRS did not result in substantial attenuation of the effect (*R^2^* = 0.012, *β* [95%CI] = -0.122 [-0.225--0.02], *p* = 0.019). Higher transcriptional risk for schizophrenia was also associated with an earlier age of onset (pT = 0.1, *R^2^* = 0.023, *β* [95%CI] = -0.22 [-0.358--0.083], *p_FDR_* = 0.02). Conversely, higher transcriptional risk for PD was significantly associated with a later age of onset (pT = 1 x 10^-4^, *R^2^* = 0.021, *β* [95%CI] = 0.221 [0.077-0.366], *p_FDR_* = 0.023), as was higher transcriptional risk for depression (pT_COLOC_ = 0.001, *R^2^* = 0.031, *β* [95%CI] = 0.391 [0.184-0.598], *p_FDR_* = 0.013).

Despite no association between transcriptional risk for PD and PD case/control status, higher transcriptional risk for PD was significantly associated with greater overall PD severity at baseline (pT = 0.05, *R^2^* = 0.026, *β* [95%CI] = 0.261 [0.105-0.417], *p_FDR_* = 0.02). Higher transcriptional risk for AD was also associated with greater PD severity at baseline (pT = Bonferroni, *R^2^* = 0.017, *β* [95%CI] = 0.136 [0.036-0.236], *p_FDR_* = 0.039). Again, higher transcriptional risk for depression and schizophrenia were associated with lower overall baseline severity (pT_DEP_ = Bonferroni; pT_SCZ_ = 1 x 10^-4^, *R^2^* range = 0.018-0.03, *β* range = -0.244--0.151, *p_FDR_* = 0.035).

Higher polygenic risk and transcriptional risk for schizophrenia were both associated with the rate of change in PD severity. However, associations were in the opposite direction, with higher polygenic risk for schizophrenia associated with a faster increase (*R^2^* = 0.024, *β* [95%CI] = 0.154 [0.053-0.255], *p_FDR_* = 0.023) but higher transcriptional risk associated with a slower increase (pT = 0.1, *R^2^* = 0.018, *β* [95%CI] = -0.196 [-0.345--0.048], *p_FDR_* = 0.037). The schizophrenia PRS and TRS showed a small nominally significantly correlation (*r* (361) = 0.133, p = 0.011). Higher polygenic risk for depression was also associated with a faster increase in PD severity (*R^2^*= 0.016, *β* [95%CI] = 0.13 [0.027-0.232], *p_FDR_* = 0.0246). There was limited evidence of effect attenuation in covariate-adjusted sensitivity analyses, with all associations remaining nominally significant when fully adjusted (**Figure 1b; Supplementary Table 11**).

#### 3.3.3 Psychiatric outcomes

Polygenic and transcriptional risk score associations with depression and anxiety in Parkinson’s at baseline can be observed in **Figure 2a**, hallucinations and apathy in Figure 2b and all psychiatric slopes in **Figure 2c**.

**Figure 2:**
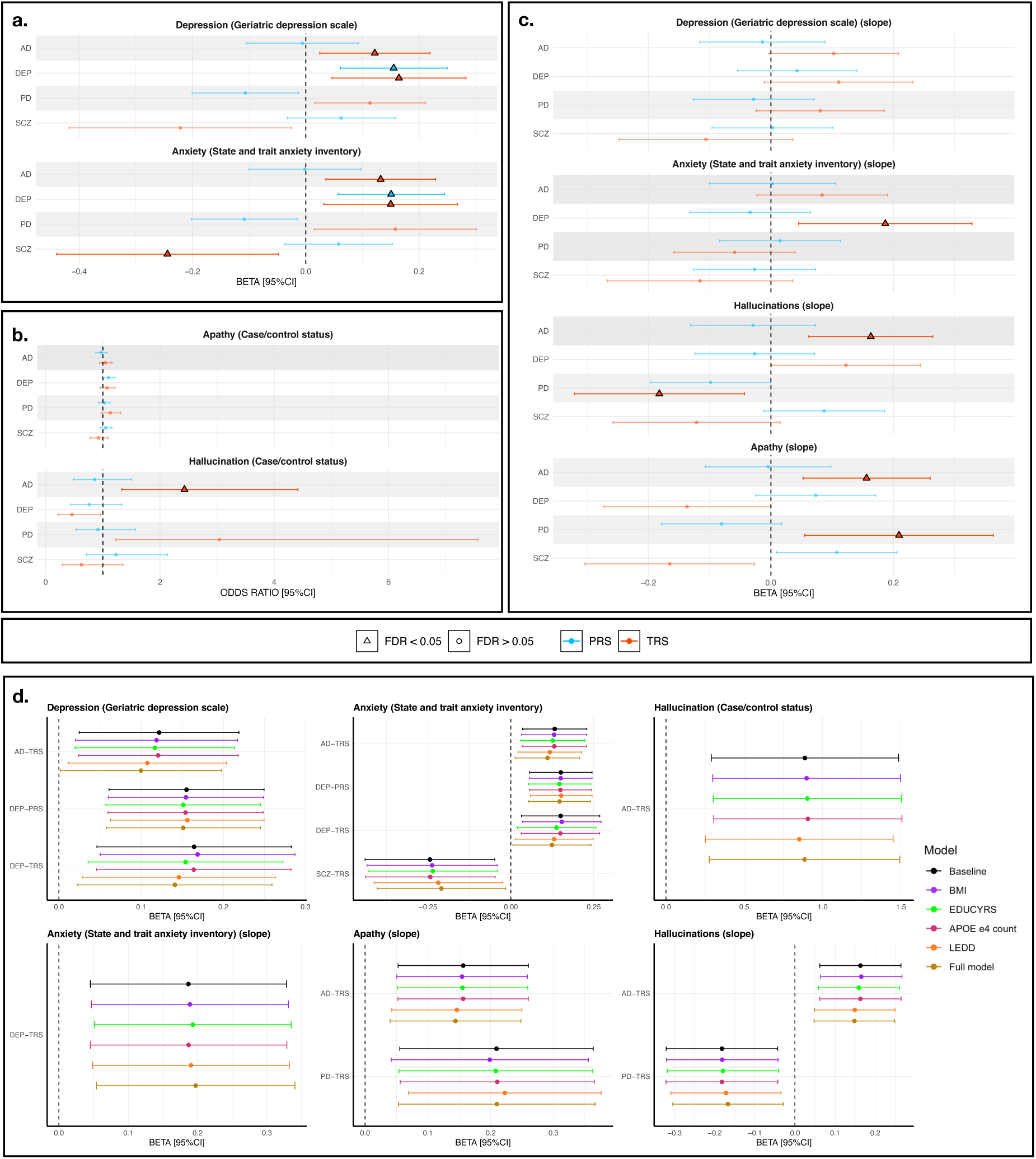
Forest plot for associations between Alzheimer’s disease (AD), depression (DEP), Parkinson’s disease (PD) and schizophrenia (SCZ) polygenic/transcriptional risk and baseline and slope psychiatric outcomes. (a) Primary analysis with baseline depression and anxiety. Point estimates for associations with p_FDR_ < 0.05 are highlighted as triangles with a black outline. Whether the estimate is from polygenic (PRS) or transcriptional risk score (TRS) is indicated in blue or orange, respectively. (b) Primary analysis with baseline apathy and hallucinations. (c) Primary analysis with the slopes of all psychiatric outcomes. (d) Covariate-adjusted sensitivity analyses for estimates with p_FDR_ < 0.05. Baseline models (e.g unadjusted models) are indicated in black, models adjusting for body mass index (BMI) in purple, years of education (EDUCYRS) in green, number of APOE e4 variants in pink, levodopa equivalent dose (LEDD) in yellow and fully adjusted models, adjusted for all covariates, in gold.

Higher polygenic risk for depression was significantly associated with higher depression and anxiety at baseline (*R^2^* range = 0.022-0.023, *β* range = 0.151-0.155 [0.061-0.249], *p_FDR_* = 0.028) as was higher transcriptional risk for depression (pT = 0.1, *R^2^* range = 0.014-0.017, *β* range = 0.15-0.164 [0.046-0.283], *p_FDR_* range = 0.033-0.046). The depression PRS and TRS were not significantly correlated (*r* (429) = 0.037, *p* = 0.442). Higher transcriptional risk for depression was also associated with a faster increase in anxiety over time (pT = 0.05, *R^2^* = 0.016, *β* [95%CI] = 0.187 [0.45-0.328], *p_FDR_* = 0.039).

Higher transcriptional risk for AD was associated with higher baseline depression and anxiety (pT_COLOC_ = Bonferroni, *R^2^*range = 0.014-0.016, *β* range = 0.122-0.132 [0.025-0.219], *p_FDR_* = 0.036-0.046). Likewise, higher transcriptional risk for AD was significantly associated with higher odds of hallucinations at baseline, but with wide confidence intervals, likely due to the low number of hallucination cases (pT_COLOC_ = 1 x 10^-6^, Nagelkerke’s pseudo-*R^2^* = 0.075, OR [95%CI] = 2.428 [1.335-4.416], *p_FDR_* = 0.023). However, an association was also observed between transcriptional risk for AD and a faster increase in hallucination symptoms over time (pT_COLOC_ = Bonferroni, *R^2^* = 0.024, *β* [95%CI] = 0.163 [0.062-0.265], *p_FDR_* = 0.023). An association was also observed between higher transcriptional risk for AD and a faster increase in apathy (pT_COLOC_ = 1 x 10^-6^, *R^2^* = 0.021, *β* [95%CI] = 0.156 [0.053-0.26], *p_FDR_* = 0.023). Conversely, higher transcriptional risk for PD was associated with a slower increase in hallucinations over time (pT = 0.05, *R^2^* = 0.017, *β* [95%CI] = 0.209 [0.055-0.363], *p_FDR_* = 0.039).

The AD transcriptional risk scores significantly associated with psychitaric symptoms did not contain genes in the *APOE* region. As such no *APOE* exclusion sensitivity analyses were performed. However, in the covariate adjusted sensitivity analyses, the association between AD transcriptional risk and baseline depression had confidence intervals with a lower bound near zero when fully adjusted (pT_COLOC_ = Bonferroni, *R^2^* = 0.009, *β* [95%CI] = 0.1 [0.002-0.198], *p* = 0.046), as did the depression TRS in association with baseline anxiety (pT = 0.1, *R^2^* = 0.009, *β* [95%CI] = 0.124 [0.006-0.242], *p* = 0.039) (**Figure 2d**), although both remained nominally significant. The remaining associations showed limited attenuation when adjusted (**Supplementary Table 12**).

#### 3.3.4 Cognitive outcomes

Associations with baseline cognitive outcomes and their slopes can be observed in Figure 3a. Covariate adjusted sensitivity analyses can be viewed in Figure 3b and Supplementary Table 13.

**Figure 3:**
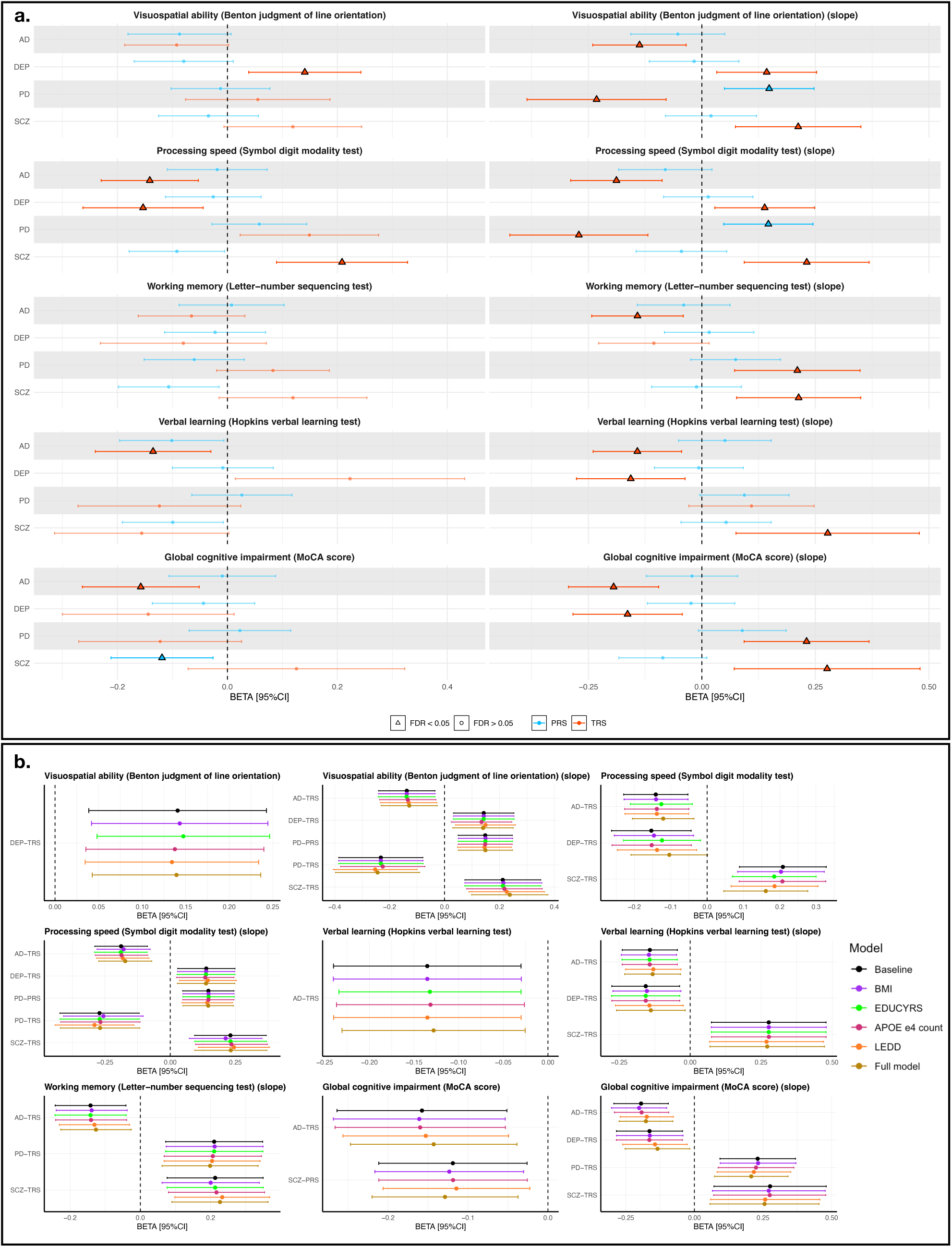
Forest plot for associations between Alzheimer’s disease (AD), depression (DEP), Parkinson’s disease (PD) and schizophrenia (SCZ) polygenic/transcriptional risk and baseline cognitive outcomes and their slopes. (a) Primary analyses. Point estimates for associations with p_FDR_ < 0.05 are highlighted as triangles with a black outline. Whether the estimate is from polygenic (PRS) or transcriptional risk score (TRS) is indicated in blue or orang,e respectively. (b) Covariate-adjusted sensitivity analyses for estimates with p_FDR_ < 0.05. Baseline models (e.g unadjusted models) are indicated in black, models adjusting for body mass index (BMI) in purple, years of education (EDUCYRS) in green, number of APOE e4 variants in pink, levodopa equivalent dose (LEDD) in yellow and fully adjusted models, adjusted for all covariates, in gold.

Neither higher polygenic nor transcriptional risk for PD were significantly associated with any cognitive outcome at baseline (p_FDR_ > 0.05). However, higher PD transcriptional risk was associated with faster decline in visuospatial ability and processing speed (pT = 0.001, *R^2^* range = 0.021-0.029, *β* range = -0.271--0.232, *p_FDR_* range = 0.013-0.023), although polygenic risk for PD showed significant associations in the opposite direction for both, with nearly identical estimates (R^2^ = 0.021, *β* [95%CI] = 0.148 [0.049-0.247], *p_FDR_* range = 0.024-0.025). The PD-PRS and TRS were not significantly correlated (*r* (429) = 0.021, *p* = 0.659). Further, higher transcriptional risk for PD was associated with slower decline in working memory and global cognitive impairment (pT_COLOC_ = 1 x 10^-6^, *R^2^* range = 0.021-0.025, *β* [95%CI] = 0.21-0.231 [0.072-0.348], *p_FDR_* = 0.02-0.025)

At baseline, higher AD transcriptional risk was associated with slower processing speed, worse verbal learning ability and worse global cognitive impairment (pT = 1 x 10^-4^, *R^2^* range = 0.014-0.019, *β* range = -0.158--0.135, *p_FDR_* = 0.02-0.042). No genes from the *APOE* region were included in the TRS score associated with global cognitive impairment. As such, no *APOE*-related sensitivity analysis was performed for these associations. Exclusion of the APOE region had a limited attenuating effect on the association with verbal learning ability (*R^2^* = 0.013, *β* [95%CI] = -0.118 [-0.213—0.022], *p_EMPIRICAL_* = 0.016). However, the association with baseline processing speed showed some attenuation following exclusion, with the upper bound of the confidence interval effectively zero (*R^2^* = 0.008, *β* [95%CI] = -0.092 [-0.183--0.001], *p_EMPIRICAL_* = 0.046).

Higher AD transcriptional risk was significant associated with faster decline in all cognitive outcomes (pT/pT_COLOC_ = Bonferroni, *R^2^* range = 0.017-0.034, *β* range = -0.194- -0.137, *p_FDR_* range = 5.866 x 10^-3^ – 0.039). Only the association with decline in verbal learning showed any attenuation in significance when removing the *APOE* region from the TRS, as this was the only score containing genes in this region, and nonetheless remained nominally significant (pT = Bonferroni, *R^2^* = 0.013, *β* [95%CI] = -0.119 [-0.219- -0.019], *p_EMPIRICAL_* = 0.021).

Higher transcriptional risk for depression was significantly associated with better visuospatial ability at baseline and slower visuospatial decline (pT_COLOC_ = Bonferroni, *R^2^* range = 0.015-0.016, *β* range = 0.141-0.143, *p_FDR_* range = 0.036-0.042). A significant association was also observed between higher transcriptional risk for depression and worse processing speed at baseline but a slower decline in processing speed over (pT = 0.1, *R^2^* = 0.015, *β* [95%CI] = -0.1531 [-0.263--0.044], *p_FDR_* = 0.035) but a slower decline in processing speed over time (pT_COLOC_ = Bonferroni, *R^2^* = 0.015, *β* [95%CI] = 0.138 [0.028-0.248], *p_FDR_* = 0.047). However, the baseline association became non-significant in the fully adjusted sensitivity model (pT = 0.1, *R^2^* = 0.007, *β* [95%CI] = -0.104 [-0.209- -0.002], *p* = 0.054). This was the only association with a cognitive outcome to show substantial attenuation in covariate-adjusted sensitivity analyses. Conversely, higher transcriptional risk for depression was associated with faster decline in verbal learning and global cognitive impairment (pT = 0.1, *R^2^* range = 0.015-0.017, *β* range = -0.164-- 0.154, *p_FDR_* = 0.031-0.039).

Higher polygenic risk for schizophrenia was significantly associated with worse global cognitive impairment at baseline (*R^2^* = 0.013, *β* [95%CI] = -0.119 [-0.212--0.026], *p_FDR_* = 0.043) while higher schizophrenia transcriptional risk was associated with faster baseline processing speed (pT = 1 x 10^-4^, *R^2^* = 0.022, *β* [95%CI] = 0.209 [0.089-0.328], *p_FDR_* = 0.011). Schizophrenia transcriptional risk was significantly associated with slower decline in all cognitive outcomes (pT range = 1 x 10^-4^ - 0.05, *R^2^* range = 0.017-0.026, *β* range = 0.213 – 0.277, *p_FDR_* range = 0.018-0.038).

## 4. Discussion

Here, we combined transcriptome-wide analysis of AD, depression, PD and schizophrenia with RNA-seq data from the Parkinson’s Progressive Markers Initiative (PPMI) to conduct the first within- and cross-trait TRS study of PD, its severity, psychiatric and cognitive symptoms, and their progression. TRS associations were compared to those from PRS.

As expected from previous work^8^, polygenic risk for PD was significantly associated with PD case/control status. However, we find no significant association with PD transcriptional risk – in contrast to previous within-trait TRS prediction of other diseases^15–19^. Nonetheless, higher transcriptional risk for PD was significantly associated with greater baseline PD severity, suggesting that while PD-TRS may not be powerful enough to differentiate cases from controls, they capture the wider burden of disease. Future work with more powerful PD GWAS and eQTL panels – resulting in improved TWAS power – may improve the downstream predictive power of PD transcriptional scores in case/control analysis.

Additionally, we found that higher transcriptional risk for AD was not only associated with greater baseline PD severity but also PD case/control status. Previous work suggests no significant genetic correlation between AD and PD^43^, yet AD-PRS can significantly predict the incidence of PD^67^. Our results suggest the contribution of AD biology to PD risk extends to profiles of AD-related gene expression, implicating shared mechanisms underlying neurodegenerative diseases.

Our findings also highlight genetic and transcriptional contributions to psychiatric symptoms in PD. Higher polygenic and transcriptional risk for depression were associated with increased baseline depression and anxiety in PD patients. Given the frequent co-morbidity of depression and anxiety and their shared genetic architecture^68^, it is not unexpected that these symptoms are associated with underlying genetic liability for depression. However, it is interesting to note that these results indicate a biological similarity between depression/anxiety as they manifest in PD and their genetic risk in the wider population. Further, results suggest their biological similarity extends to profiles of gene expression. As depression PRS and TRS were not significantly correlated, they appear to represent unique biological contributions to anxiety/depression. Given the phenotypic and genetic heterogeneity of depression^69^, novel insights into pathophysiology might be derived by comparing the symptomatic profiles of individuals with high depression PRS to those with high depression TRS in future, depression focussed work. Additionally, higher transcriptional risk for depression was associated with a more rapid increase in anxiety over time. Since this association was not identified with depression PRS, it is plausible that while polygenic risk contributes to a general baseline predisposition for higher depression and anxiety, TRS better capture the acute disease biology driving progression, being closer to the phenotype on the omics cascade^70^.

Higher transcriptional risk for AD was also associated with baseline depression and anxiety in individuals with PD, as well as with the presence of hallucinations at baseline and a more rapid increase in apathy and hallucinations over time. Previous work suggests there is limited genetic overlap between AD and depression^45^, and there is evidence that depression and anxiety symptoms in AD are instead linked to AD-specific neurodegenerative processes, such as cortical atrophy, elevated phosphorylated tau protein and lower levels of cerebrospinal fluid amyloid-beta^71^. Apathy is also common in AD, observed in approximately 50% of patients, while hallucinations are observed in approximately 16%^72^. That associations between transcriptional risk for AD and these symptoms are observed in PD again suggests shared biological mechanisms across neurodegenerative diseases. Further, as no associations were identified with AD-PRS, our findings suggest that AD-specific biological contributions to psychiatric symptoms are better captured through profiles of gene expression, at least within PD. It may therefore be of interest to evaluate AD-TRS associations with psychiatric symptoms in AD itself for comparison in future work.

Hallucinations are a suggested side-effect of anti-Parkinson’s treatments such as levodopa and dopamine replacement therapy due to their hypersensitisation of dopaminergic neurons in the substantia nigra^73^ – the primary area of neurodegeneration in PD^74^. However, hallucinations are also observed in PD patients pre-medication and have been linked to the progression of PD-related structural brain changes^75^. Here, we find that controlling for levodopa equivalent dose had no effect on any hallucination-related associations, suggesting they are not confounded by pharmaceutical side-effects. However, interpretation of the baseline association warrants caution given the small number of cases and the breadth of hallucination severity included in the case definition due to the dichotomisation of an ordinal outcome.

Among cognitive outcomes, transcriptional risk for AD was associated with faster decline in all cognitive domains, as well as worse baseline processing speed, verbal learning and global cognitive impairment. Although previous work has linked the *APOEε4* risk variants to PD-related cognitive decline^66,76^, we find that controlling for the number of *APOEε4* variants did not affect associations, while excluding genes in the *APOE* region from the AD-TRS, where they were originally included, only attenuated the baseline association with verbal learning. This suggests that associations are driven by genome-wide profiles of gene expression linked to AD, as opposed to the expression of genes in the *APOE* region alone. As with psychiatric symptoms, we find no associations between AD-PRS and cognitive outcomes. Such results emphasise the potential strength of transcriptional risk scores to identify individuals at risk of cognitive decline in PD, enabling earlier interventions.

In contrast to AD, we observed that transcriptional risk for schizophrenia was associated with a slower decline across all cognitive outcomes. Similar protective effects were observed for PD case/control status, lower disease severity, a slower increase in PD severity over time and lower baseline anxiety. Schizophrenia is associated with dopaminergic hyperactivity and the majority of anti-psychotics act as antagonists for dopamine D2 receptors^77^. Anti-psychotic use is the second most common cause of Parkinsonism after PD itself, with motor symptoms persisting even after medication use ceases^78^. It is possible that individuals with higher gene expression related to schizophrenia have greater dopaminergic activity, which may in turn protect against the neurodegeneration of striatal dopamine neurons seen in PD. However, it is worth noting that polygenic risk for schizophrenia is associated worse cognition in individuals with schizophrenia^79^. As such, further work is required to better understand the effects of schizophrenia-related gene expression in PD specifically, with the potential to uncover novel mechanisms of neuroprotection.

The results were not without their contradictions. Converse to TRS associations, higher schizophrenia polygenic risk was associated with a faster increase in PD severity over time as well as worse global cognitive impairment at baseline. Similarly, among cognitive outcomes, depression transcriptional risk was associated with a faster decline in some domains – including overall cognitive impairment – but a slower decline in others. Similar patterns were observed for cognitive slope outcomes and PD transcriptional risk. In some cases, polygenic and transcriptional risk for PD were both significantly associated, but in opposite directions. These findings might suggest that PRS and TRS capture different aspects of disease biology, highlighting the complexity of omic-based prediction and the importance of comparing associations across omic layers. Future work might incorporate omic-based risk scores from proteomic and metabolic data for further granularity. Both PRS and TRS utilise GWAS summary statistics to obtain genetic weights, though the TWAS-derived TRS weights specifically capture genetic effects linked to altered gene expression. As noted in work utilising TWAS-imputed gene expression, imputed TRS are directionally concordant with PRS, but explain a smaller proportion of the variance and overall heritability^80^. However, the TRS calculated here use observed gene expression levels in the target sample, which may be subject to non-genetic influences, such as environmental exposures or the heterogeneity of disease pathology. This may also help explain differences in the direction of associations between PRS and TRS.

Nonetheless, it is notable that the vast majority of significant associations were identified using TRS not PRS. This study intentionally restricted the sample to individuals in PPMI with both genetic and transcriptomic data available, allowing us to compare associations between the different methods in identically sized samples. Such results imply that TRS may have greater power for detection and can identify biologically driven associations in PD relevant phenotypes missed by PRS.

However, these results must be viewed in light of several limitations. As in previous TRS work^15,19^, this study was conducted in a single cohort. Replication is a crucial future step here given several contradictory results. Further, samples were restricted to individuals of European ancestry only. As is well noted, polygenic scores from European ancestry GWAS have limited portability to other ancestries^81^. As TRS are in their infancy compared to PRS, evaluation of the cross-ancestry portability of TRS calculated using observed gene expression have yet been conducted. However, TRS calculated using imputed gene expression have shown promise^82^. Future TRS work should explore predictive ability of TRS calculated using observed gene expression across ancestry-diverse target samples. Sample size may also be a limiting factor. Although the PPMI RNA-seq sample is comparable to widely used gene expression resources such GTEx^83^, the sample size is small for polygenic scoring compared to available genotyped resources like the UK Biobank^84^. Power differences may partially explain why most associations were detected with TRS and not PRS. Future comparative work is required. Similarly, here we use RNA-seq data derived from blood. It is possible that prediction would improve using RNA-seq quantified in brain tissues, given its primacy in PD pathology and thus greater proximity to the disease. The comparison of TRS prediction using RNA-seq data from different tissues is an important future step likely to provide further insight into underlying biology. Finally, the clumping and thresholding TRS approach used here is likely to lead to inflated *R^2^* values due to overfitting. Although we guard against false positives using permutation tests to estimate empirical *p*-values and use a further FDR correction, external validation must once again be emphasised.

In conclusion, this study represents the first transcriptional risk score study of PD, its symptoms and their progression. Findings suggest that TRS can detect cross-trait biological contributions to PD and its symptoms not detected by PRS and may prove of particular use for identifying individuals at risk of progression.

## 5 Acknowledgements

LG is funded by the King’s College London DRIVE-Health Centre for Doctoral Training and the Perron Institute for Neurological and Translational Science. PP is funded by Alzheimer’s Research UK. OP is supported by a Sir Henry Wellcome Postdoctoral Fellowship [222811/Z/21/Z]. SK and ALP is funded by MSWA and the Perron Institute. AI is funded by South London and Maudsley NHS Foundation Trust, MND Scotland, Motor Neurone Disease Association, National Institute for Health and Care Research, Spastic Paraplegia Foundation, Rosetrees Trust, Darby Rimmer MND Foundation, the Medical Research Council (UKRI) and Alzheimer’s Research UK. HM is funded by an MND-Scotland/Motor Neurone Disease Association (MNDA) Pre-Fellowship Grant. This paper represents independent research part-funded by the NIHR Maudsley Biomedical Research Centre at South London and Maudsley NHS Foundation Trust and King’s College London. The views expressed are those of the author(s) and not necessarily those of the NIHR or the Department of Health and Social Care. The authors thank Million Veteran Program (MVP) staff, researchers, and volunteers, who have contributed to MVP, and especially participants who previously served their country in the military and now generously agreed to enrol in the study. (See https://www.research.va.gov/mvp/ for more details). The citation for MVP is Gaziano, J.M. et al. Million Veteran Program: A mega-biobank to study genetic influences on health and disease. J Clin Epidemiol 70, 214-23 (2016). This research is based on data from the Million Veteran Program, Office of Research and Development, Veterans Health Administration, and was supported by the Veterans Administration (VA) Million Veteran Program (MVP) award #000. We want to acknowledge the participants of the FinnGen study and the FinnGen research team. We also thank and acknowledge the contribution and use of the CREATE high-performance computing cluster at King’s College London (King’s College London. (2022). King’s Computational Research, Engineering and Technology Environment (CREATE)). For the purposes of open access, the author has applied a Creative Commons Attribution (CC BY) license to any Accepted Author Manuscript version arising from this submission.

## 6. Data availability

Data used in the preparation of this article were obtained on January 29^th^ 2024 from the LONI Image & Data Archive (LONI IDA) (https://ida.loni.usc.edu/home/projectPage.jsp?project=PPMI) and is also available through the Parkinson’s Progression Markers Initiative (PPMI) database (https://www.ppmi-info.org/access-data-specimens/download-data), RRID:SCR_006431. For up-to-date information on the study, visit http://www.ppmi-info.org. PPMI – a public-private partnership – is funded by the Michael J. Fox Foundation for Parkinson’s Research and funding partners, including 4D Pharma, Abbvie, AcureX, Allergan, Amathus Therapeutics, Aligning Science Across Parkinson’s, AskBio, Avid Radiopharmaceuticals, BIAL, BioArctic, Biogen, Biohaven, BioLegend, BlueRock Therapeutics, Bristol-Myers Squibb, Calico Labs, Capsida Biotherapeutics, Celgene, Cerevel Therapeutics, Coave Therapeutics, DaCapo Brainscience, Denali, Edmond J. Safra Foundation, Eli Lilly, Gain Therapeutics, GE HealthCare, Genentech, GSK, Golub Capital, Handl Therapeutics, Insitro, Jazz Pharmaceuticals, Johnson & Johnson Innovative Medicine, Lundbeck, Merck, Meso Scale Discovery, Mission Therapeutics, Neurocrine Biosciences, Neuron23, Neuropore, Pfizer, Piramal, Prevail Therapeutics, Roche, Sanofi, Servier, Sun Pharma Advanced Research Company, Takeda, Teva, UCB, Vanqua Bio, Verily, Voyager v. 25MAR2024 Therapeutics, the Weston Family Foundation and Yumanity Therapeutics.

Summary statistics from FinnGen are publicly available via their website (https://www.finngen.fi/en/access_results). Summary statistics for depression and schizophrenia from the Psychiatric Genomics Consortium (PGC) are publicly available on their website (https://pgc.unc.edu/for-researchers/download-results/). Summary statistics from the Million Veteran Project via dbGaP request (https://www.ncbi.nlm.nih.gov/projects/gap/cgi-bin/study.cgi?studyid=phs001672.v1.p1). eQTL datasets used by FUSION for TWAS are available through the FUSION website (http://gusevlab.org/projects/fusion/#download-pre-computed-predictive-models).

